# A distinct class of pan-cancer susceptibility genes revealed by alternative polyadenylation transcriptome-wide association study

**DOI:** 10.1101/2023.02.28.23286554

**Authors:** Hui Chen, Zeyang Wang, Jia Wang, Wenyan Chen, Xuelian Ma, Xudong Zou, Mireya Plass, Cheng Lian, Ting Ni, Gong-Hong Wei, Wei Li, Lin Deng, Lei Li

## Abstract

Alternative polyadenylation (APA) plays an important role in cancer initiation and progression; however, current genome- and transcriptome-wide association studies (GWAS and TWAS, respectively) mostly ignore APA when identifying putative cancer susceptibility genes. Here, we performed a pan-cancer 3′untranslated region (UTR) APA TWAS (3′aTWAS) by integrating 80 well-powered (*n*>50,000) GWAS datasets across 23 major cancer types with APA quantification from 17,330 RNA sequencing samples across 49 tissue types and 949 individuals. We found that genetic variants associated with APA represent around 24.4% of cancer GWAS variants and are more likely to be causal variants explaining a large portion of cancer heritability. We further identified 413 significant APA-linked cancer susceptibility genes. Of these, 77.4% have been overlooked by traditional expression- and splicing-studies, given that APA may regulate translation, protein localization, and protein–protein interactions independent of the expression level of the genes or splicing isoforms. As proof of principle validation, modulation of four novel APA-linked breast-cancer susceptibility genes significantly altered cancer cell proliferation. Our study highlights the significant role of APA in discovering new cancer susceptibility genes and provides a strong foundational framework for enhancing our understanding of the etiology underlying human cancers.

## Introduction

Genome-wide association studies (GWAS) have identified hundreds of single-nucleotide polymorphisms (SNPs) associated with increased risks of major human cancers^1, 2^, including breast^3^, prostate^4^, colorectal^5^, and ovarian cancer^6^. In prostate cancer, for example, a highly heritable disease with 58% risk due to genetic factors, over 140 prostate-cancer-risk variants have been identified, explaining approximately one-third of familial risk for disease^7^. Improving our understanding of inherited cancer-risk-associated SNPs could provide new opportunities to elucidate the mechanisms of tumorigenesis. However, more than 90% of these variants are mapped to noncoding regions of the human genome^8^, posing a significant challenge for their functional interpretation in regard to disease development, progression, and response to therapy.

Molecular quantitative trait locus (xQTL) analysis is a crucial step toward better understanding the effects of noncoding genetic variants on genes, pathways, and mechanisms of action, serving as an essential intermediate link between genotype and disease phenotype^9–11^. Many molecular phenotypes derived from RNA sequencing (RNA-seq), such as gene expression and alternative splicing, have been used to discover disease-risk genes in population-scale xQTL studies^10, 12^. Those genetic variants showing strong associations with the aforementioned molecular phenotypes are referred to as expression QTL (eQTL) or splicing QTL (sQTL). Such xQTLs can be highly informative; however, the effects of the numerous disease-associated noncoding variants remain unexplained^9, 13^.

Alternative polyadenylation (APA) has emerged as a new paradigm of post-transcriptional regulation for human genes^14–16^. That is, by employing different poly(A) sites, genes can either shorten or extend 3′untranslated regions (UTRs) containing *cis*-regulatory elements, such as binding sites for microRNAs or RNA-binding protein (RBP)^17^. APA can affect target gene translation, as well as localization and protein–protein interactions of its gene product, independent of mRNA expression level or splicing^14^. Consequently, the diverse landscape of polyadenylation can significantly impact both normal development and disease progression^18, 19^. In particular, individual genetic variants associated with APA have been linked to several cancers. For example, rs78378222 in the 3′UTR of *TP53* alters the canonical polyadenylation signal from AATAAA to AATACA; this impairs 3′-end processing of *TP53* mRNA, altering susceptibility to multiple cancers, including cutaneous basal cell carcinoma, prostate cancer, glioma, and colorectal adenoma^20^. In our previous study, we described the first atlas of genetic variants associated with APA (3′aQTL)^21, 22^, which can explain approximately 16.1% of 15 human diseases and traits, not including cancers. Therefore, the prevalence and functions of SNPs associated with APA for major human cancer types remain largely unknown.

In this study, we performed the first large-scale and systematic analysis assessing the genetic effects of APA on 25 cancer types in 49 human tissues. Our results show that widespread genetic pleiotropy and cancer type, rather than genetic ancestry, primarily contributes to cancer heritability. We further performed colocalization and transcriptome-wide association studies (TWAS) and identified 413 cancer susceptibility genes predicted to modulate cancer risk via APA, 77.4% of which are independent of gene expression and splicing. In addition, we experimentally validated four of these APA-associated risk genes linked to breast cancer, showing that interference of these genes significantly disturbed cancer cell proliferation. Lastly, we have constructed a publicly available database with a user-friendly hub (http://bioinfo.szbl.ac.cn/TCGD/index.php) for use by the research community.

## Results

### Widespread genetic pleiotropy across multiple cancer types

To comprehensively characterize the genetic effects of APA on human cancers, we compiled a large collection of 438 GWAS summary statistics from manually curated published studies and public cohorts, including the National Human Genome Research Institute–European Bioinformatics Institute (NHGRI–EBI) GWAS Catalog (release 2021/01)^23^, the UK Biobank release 2 cohort (UKB2; release 2018/03)^24^, the Japanese ENcyclopedia of GEnetic associations by Riken (JENGER), and the FinnGen consortium (release 2021/05). After filtering, summary statistics from 80 reasonably powered GWASs (*n* > 50,000) across 25 cancer types were retained for our analysis (Fig. 1a); the median sample size across these studies was 180,856 individuals. We then extracted all leading SNPs and found they tend to have large effect sizes compared to the whole genome (*P* = 1.07 × 10^-39^, Wilcoxon rank–sum test) (Fig. 1b and Fig. S1). We further analyzed their functional annotations to determine the positional distribution of leading SNPs and found 96.25% of GWAS lead SNPs were in noncoding regions, and 14.1% of them were located in 3′UTR and downstream regions (20kb) (Fig. 1c and Fig. S2), such as prostate cancer SNP rs4245739 in the 3′UTR of *MDM4,* which encodes a regulator of p53, and breast cancer SNP rs763121 in the 3′UTR of *DDX17,* which encodes the DEAD-box helicase 17. We also validated our results by performing a fine-mapping analysis, which estimates the probability of a genetic variant being causal via conditioning on all associated variants in each genomic region^25^, on these cancer GWAS data. Results show that fine-mapped SNPs have similar patterns, indicating a strong enrichment for cancer-associated variants in noncoding regions.

**Figure 1.**
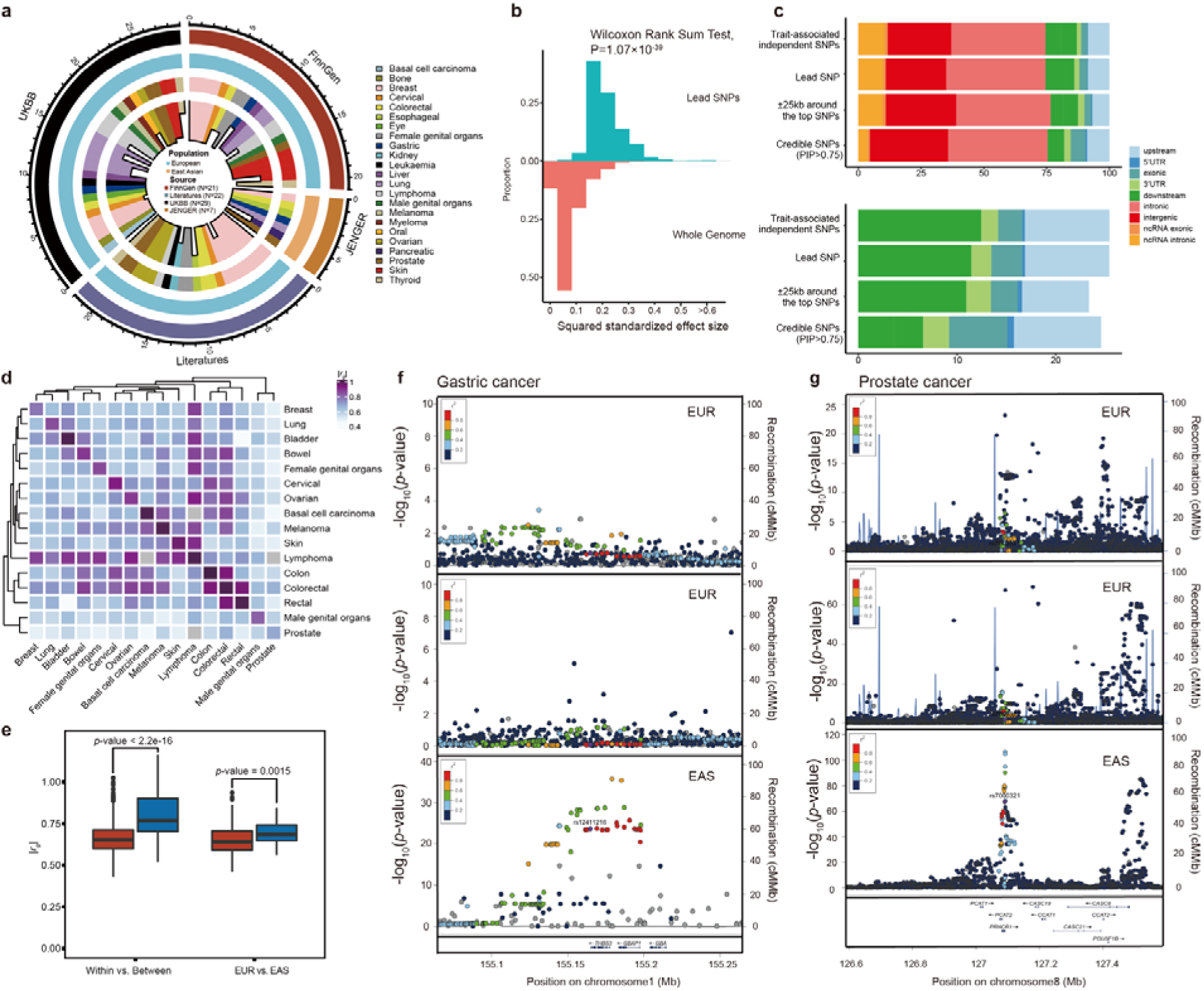
Overview of collected cancer-related genome-wide association study (GWAS) summary statistics. **a.** Atlas of genetic influences identified from 80 GWAS samples across 23 human cancer types. The inner histogram represents the distribution of leading single-nucleotide polymorphisms (SNPs) from each GWAS summary data. The third circle highlights the different cancer types indicated by different colors. The second circle represents the different populations including European and Asian people indicated by blue and brown colors. The outside circle highlights the data sources as indicated by different colors. **b.** Proportional histogram of squared standardized effect sizes for the leading SNPs, with minor allele frequency (MAF) > 0.01. **c**. Positional distribution of SNPs with functional consequences. **d.** Genetic correlation between cancers. Heatmap reflects the average *r_g_* for all nominally significant (*P* < 0.05) trait pairs. **e.** Comparisons of the genetic correlations within cancers *vs*. between cancers and in Europeans *vs*. Asians. Wilcoxon test *P*-value was shown. **F.** LocusZoom plot showing the significant loci associated with gastric and prostate cancer in Europeans and Asians; for example, rs12411316 in gastric cancer is only significant in the Asian population. EAS, East Asian; EUR, European.

To elucidate the degree of shared genetic architecture across different cancer types, we calculated heritability estimates from genome-wide summary statistics, using linkage disequilibrium score regression (LDSC)^26^, and found that heritability estimates vary across different cancer types. Lymphoma has the lowest average heritability score (*h*^2^ = 0.0017) with 95% confidence interval (CI): 0.0004–0.0030, whereas breast cancer has the highest average heritability score (*h^2^ =* 0.17), with 95% CI: 0.1637–0.1835 (Fig. 1d). Our estimations are well consistent with those in previous studies^5,27^. We further performed pairwise genetic correlation (*r_g_*) analysis and found that 22.09% of cancer pairs are significantly correlated at a Bonferroni-corrected threshold of *P* = 0.05. The average cancer correlation is 0.64. We also observed that biologically related cancers tend to cluster together, for example, basal cell carcinoma clusters with melanoma and skin cancers, whereas colon and rectal cancers cluster together. We then compared the genetic correlation between and within cancers and observed significantly higher genetic correlations within cancer types (average |*r_g_*| = 0.76) than between cancer types (average |*r_g_*| = 0.64) (Fig. 1e). Because genetic ancestry can also contribute to cancer heterogeneity^2^. Therefore, we analyzed the genetic correlation between different populations for each cancer type. The results reveal a weak genetic difference between within-European and within-Asian samples (Fig. 1f), suggesting that cancer type, rather than genetic ancestry, primarily contributes to cancer heritability. Taken together, these data show that 14.1% of Cancer GWAS SNPs are in 3′UTR and gene downstream regions, and human cancer samples have widespread genetic pleiotropy across multiple cancer types.

### 3′aQTL explains a large portion of cancer heritability

To assess the role of APA regulatory variants in human cancer and quantify the effect of 3′aQTL enrichment on cancer heritability, we performed 3′aQTL analysis^28^ and applied stratified (S)-LDSC^29, 30^. To the best of our knowledge, this study represents the largest 3′aQTL study of human cancers. We found that 3′aQTLs are significantly enriched in most cancer types, with a fold enrichment comparable to that of sQTLs and eQTLs, on average, across traits. We further calculated the relative contributions of eQTL, sQTL, and 3′aQTL to cancer heritability, with a false-discovery rate (FDR) < 5%. We found that 3′aQTLs can explain a median 7.5% of heritability, compared to 12.1% for sQTLs and 22.5% for eQTLs (Fig. 2a, b). Expanding our S-LDSC analysis revealed that 3′aQTLs in whole blood and adipose tissues are enriched for associations with multiple cancer types, including cancers of the skin, breast, prostate, and male genital organs, as well as basal cell carcinoma (Fig. 2c).

**Figure 2.**
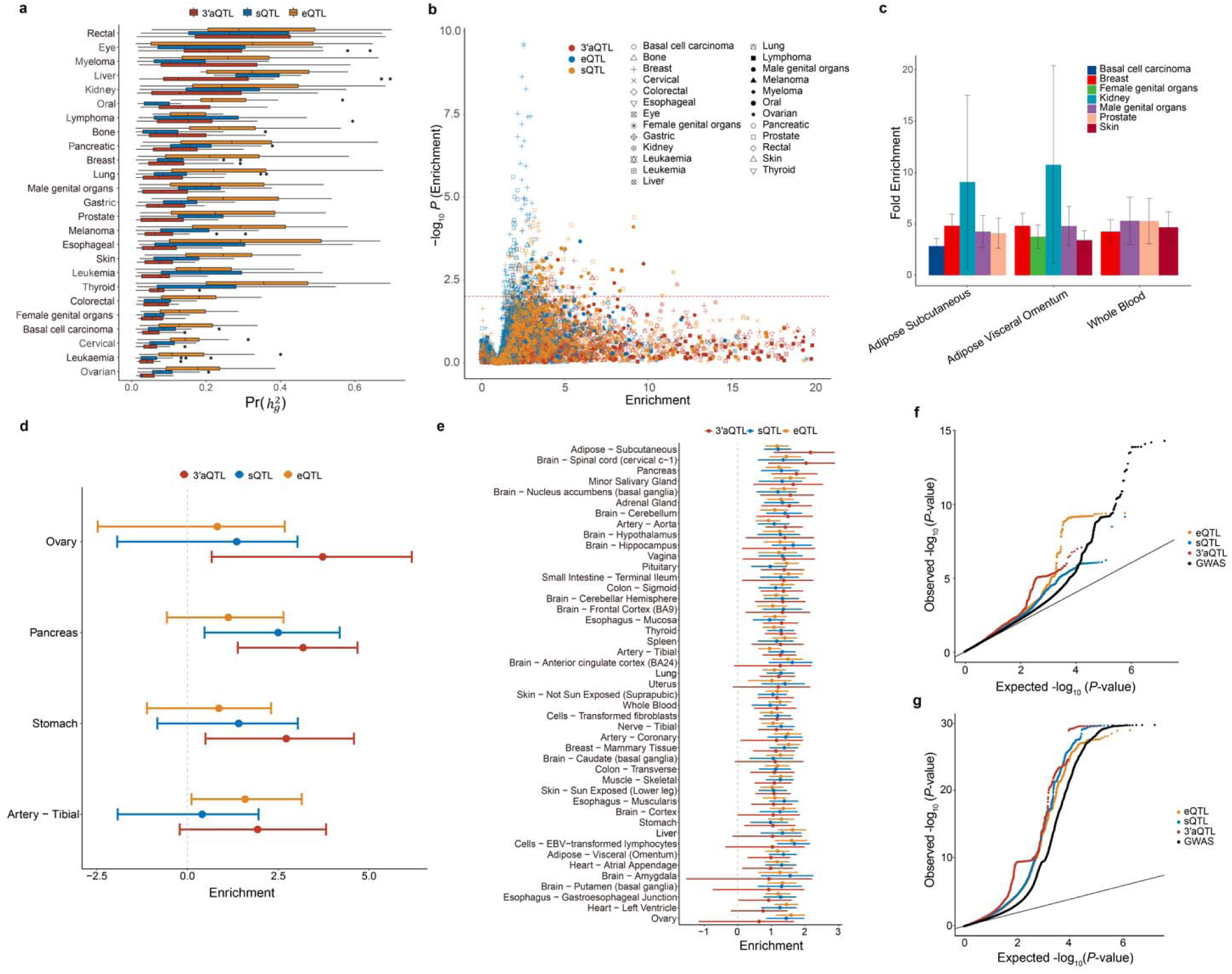
Integrated analysis of xQTL enrichment for heritability of human cancer traits. **a**. Partitioned heritability determined by calculating the ratio of phenotypic heritability (*x*-axis) attributable to expression quantitative trait loci (eQTLs), splicing QTLs (sQTLs), and alternative polyadenylation (APA) QTLs (3′aQTLs) relative to aggregate SNPs (*h*_*XQTL*_^2^/*h*_*SNP*_^2^) for different cancer types (*y*-axis). **b**. Summary of GWAS heritability enrichment for cancer traits (with the largest sample and case number) on the baseline linkage disequilibrium (LD) model. Dashed line shows the significant threshold at *P* < 0.05. **c.** Heritability enrichment in adipose-related and whole-blood tissues for multiple cancer types. The error bars represent the standard error. **d,e**. Tissues with larger enrichment of 3′aQTLs than of eQTLs or sQTLs for (d) endometrial cancer and (e) breast cancer. The effect sizes were estimated using functional genome-wide association analysis, which quantifies the enrichment of xQTLs in trait-associated variants. The estimated lower and upper bound 95% confidence intervals (CIs) for the effect sizes are also shown. **f,g.** Quantile–quantile plots of GWAS *P*-values for (d) endometrial cancer and (e) breast cancer; 3′aQTLs, eQTLs, and sQTLs are shown in comparison with genome-wide SNPs. GWAS SNPs were binarily annotated using 3′aQTLs, eQTLs, and sQTLs, with *P* < 1×10^-4^.

We next evaluated the enrichment of xQTLs by performing a functional genome-wide association study (*fgwas*) ^31^ and observed substantial 3′aQTL enrichment for multiple cancer associations. Notably, although the effects of eQTLs were found to be more significant than those of 3′aQTLs for 13.9% of tissue–cancer pairs examined, we identified increased enrichment of 3′aQTLs within 7.7% of pairs. Interestingly, 3′aQTLs displayed increased enrichment relative to eQTLs across several biologically relevant tissue–cancer pairs, such as the ovary for endometrial cancer (Enrichment=3.72, Fig. 2d) and adipose tissue for breast cancer (Fig. 2e). Quantile–quantile plots (QQ-Plots) of GWAS *P*-values for xQTLs and genome-wide SNPs further validate these results (Fig. 2f, g). Thus, our findings suggest that 3′aQTLs can explain a significant portion of cancer heritability.

### 3′aQTLs colocalize with cancer variants and are largely independent of gene expression and splicing QTLs

To systematically identify the causal cancer GWAS SNPs shared with 3′aQTLs, we performed colocalization analysis on 49 human tissues using *coloc*^32^ and SMR-HEIDI^33^. There are 62 cancer GWAS samples across 20 cancer types colocalized with at least one type of molecular QTLs (Fig. 3). We identified 913 eQTLs, 692 sQTLs, and 473 3′aQTLs that colocalize with cancer variants. Notably, a large proportion of eQTL colocalization events in long-noncoding RNAs were observed for several cancers, including colorectal cancer and lymphoma, whereas other cancer samples, such as melanoma and basal cell carcinoma, contain a high proportion of 3′aQTL colocalization events in pseudogenes (Fig. 3b). These results suggest that lncRNAs and pseudogenes may play an essential role in cancer progression.

**Figure 3.**
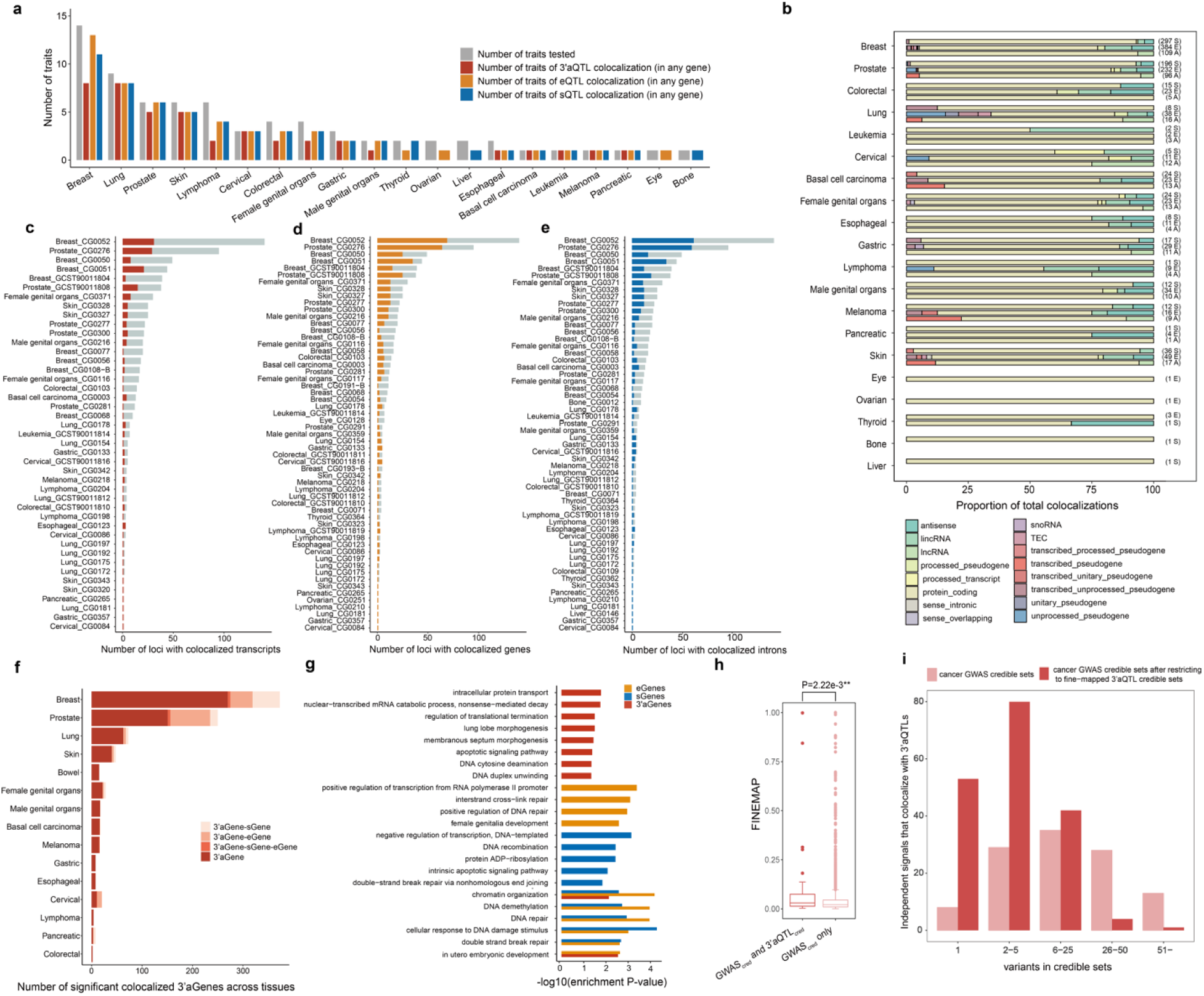
Colocalization identifies trait-associated genes. **a.** The number of specific traits for different cancer types that colocalize with 3′aQTLs, sQTLs, and eQTLs, indicated by the red, orange, and blue bars, respectively. **b.** Frequency of significant colocalization events within each gene biotype, collapsed across tissues (feature–GWAS combinations). GWAS data are grouped by general cancer categories on the *y*-axis. A missing bar indicates there was no colocalization for that given trait category and QTL type. The numbers at the right of each bar indicate the total number of significant colocalization events (E, eQTL; S, sQTL; A, 3′aQTL). **c–e.** The proportion of loci with a GWAS-significant variant that colocalizes with at least one gene expression (c), splicing (d), or 3IR uIe (e) event. Across traits, a median of 49.6%, 35.0%, and 24.4% of GWAS loci colocalize with eQTLs, sQTLs, and 3′aQTLs, Respectively. **f.** Barplot shows that **7**8.5% of 3′aQTL-colocalized cancer risk loci are specific for a 3′aQTL, without a corresponding EQTL or sQTL. **g.** Pathway analysis of 3′aGenes, sGenes, and eGenes for breast cancer. The size of the circles represents the number of genes enriched in each pathway, and the color indicates significance of enrichment. **h.** Distribution of cancer GWAS causal posterior probabilities for all fine-mapped GWAS variants (95% credible set in GWAS, GWAS_cred_) that are also fine-mapped 3′aQTL variants (3′aQTL_cred_) *vs*. GWAS_cred_ variants only. Mann–Whitney *P*-value is shown. **i.** Integration of cancer-credible GWAS variants with credible sets from colocalizing 3′aQTLs increases fine-mapping resolution. Bar plot shows the number of independent signals identified as candidate causal variants before and after restricting for QTL variants.

We then analyzed the proportion of cancer variants that can be explained by these molecular QTLs and found that 3′aQTLs, eQTLs, and sQTLs can explain a median 24.4%, 49.6%, and 35.5% of cancer variants, respectively (Fig. 3c–e). Further analysis of the 3′aQTL-colocalized genes (3′aGenes) relative to eQTL and sQTL colocalized genes (eGenes and sGenes, respectively) revealed that cancer-associated 3′aGenes are largely distinct from the others. That is, an average of 78.5% 3′aGenes are not associated with matched eQTLs or sQTLs among the analyzed tissue–cancer pairs (Fig. 3f). For example, we identified 3′aQTLs in the caspase 8 (*CASP8)* gene that strongly colocalize with GWAS variants across multiple cancers, including skin cancer (PPH_4_ = 0.961), breast cancer (PPH_4_ = 0.973), and basal cell carcinoma (PPH_4_ = 0.915), but not with eQTLs or sQTLs. Pathway analysis also revealed that 3′aGenes are enriched in several distinct pathways, such as intracellular protein transport, which contributes to the regulation of energy consumption and metabolism during cancer progression, and nonsense-mediated decay (Fig. 3g). In contrast, eGenes and sGenes are enriched in DNA damage repair, chromatin organization, and DNA demethylation pathways. Thus, our data suggest that 3′aQTLs colocalize with cancer variants, and a large proportion of these 3′aQTLs occur independently of gene expression and splicing QTLs.

To further investigate whether 3′aQTLs were also enriched at causal risk variants, we performed fine-mapping on colocalized 3′aQTLs and compared them with cancer variants within a 95% credible set. We found that cancer variants at 3′aQTLs have a significantly higher posterior inclusion probability than those not at 3′aQTLs (Mann–Whitney, *P* = 2.22 × 10^-3^; Fig. 3h). Furthermore, we found that inclusion of fine-mapped 3′aQTLs significantly increased the genetic resolution of cancer-credible sets, resulting in the identification of 133 cancer-credible sets with ≤5 variants compared to only 37 sets when 3′aQTLs are not considered (Fig. 3i). These results suggest that the identified 3′aQTLs likely contain causal cancer-risk variants, and in total, our above findings indicate that 3′aGenes genes are largely distinct from eGenes and sGenes across many cancer types.

### APA transcriptome-wide association analysis reveals novel cancer susceptibility genes

To systematically identify and prioritize candidate APA genes associated with human cancers, we performed a multi-tissue 3′UTR APA transcriptome-wide association study (3′aTWAS) by integrating our curated cancer GWAS summary statistics and transcriptome panels (gene expression, splicing, and APA) from 49 tissues. From this analysis, we obtained 92,062 tissue-specific prediction models (Fig. S2a), including 20,613 APA events. We then evaluated the prediction accuracy by calculating the correlation between predicted and observed 3′UTR usage and further normalized by heritability score. Results show that the average in-sample prediction accuracy is 80.9%, and the accuracy score is consistent with previous TWAS studies^34, 35^. These data indicate that our prediction model has successfully captured cis-regulated APA usage. Moreover, the number of 3′aTWAS predictive models is highly correlated with the sample sizes of the reference panels (Spearman correlation R = 0.83, *P* = 1.46 × 10^-13^; Fig. S2b).

We further applied our prediction model to cancer GWAS data and identified 335 APA-linked cancer susceptibility events (FDR < 0.05), most of which were overlooked by conventional TWAS analyses (Fig. 4a-d). Notably, our 3′aTWAS identified multiple known and novel cancer susceptibility genes, such as cathepsin F (*CTSF),* which is significantly associated with breast cancer in breast mammary tissue (Z = -3.71, *P* = 2.07 × 10^-4^, FDR = 0.014). This finding suggests that APA of *CTSF*, rather than gene expression or splicing of the *CTSF* transcript, mediates breast cancer risk. In addition, APA of ADP ribosylation factor-like GTPase 17B (*ARL17B)* was found to be associated with cervical cancer in the female genital organs. Nicolin 1 (*NICN1)* is another example that highlights how APA may independently contribute to cancer risk by regulating the subcellular localization of mRNA isoforms. That is, we observed a positive Z-score for *NICN1* in prostate cancer, indicating that extended 3′UTR usage of *NICN1* promotes prostate cancer risk. This distal APA isoform of *NICN1* includes inverted Alu repeats that lead to its nuclear retention, whereas in contrast, the proximal APA isoform lacking these sequences is located within cytoplasm^36^.

**Figure 4.**
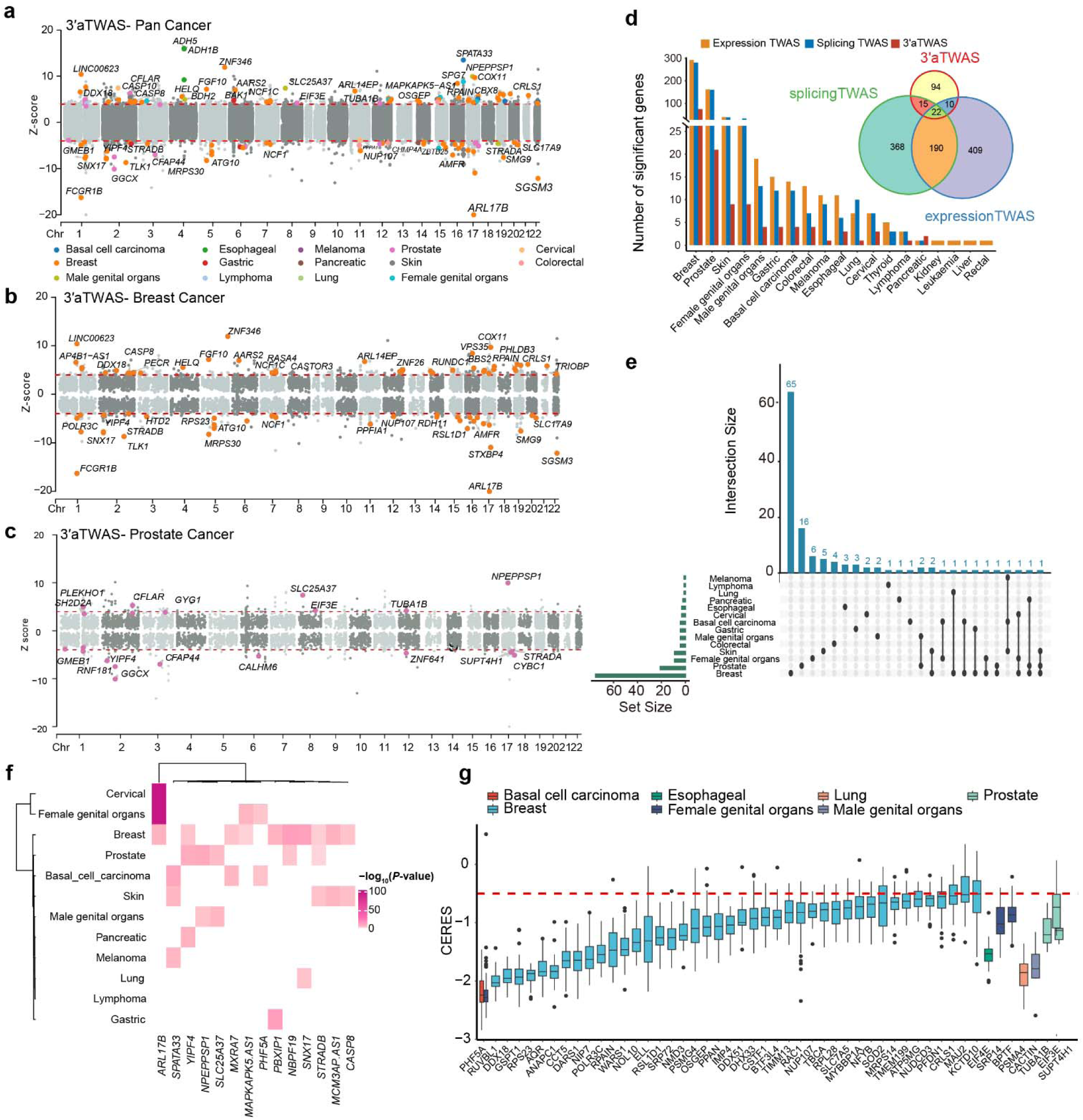
Transcriptome-wide association study (TWAS) results. **a-c**. Manhattan plot of 3′aTWAS nominating the susceptibility genes in (a) pan-cancer, (b)breast cancer and (c) prostate cancer, respectively. Colored points represent significant 3′aTWAS associations at FDR<0.05. **d.** Bar plots showing the number of significant genes detected by 3′aTWAS, with a false-discovery rate (FDR) < 0.05, for different cancers in the most relevant tissues. Venn plot shows the intersection of significant cancer genes identified by 3′aTWAS (FDR <0.05) with genes identified by expression and splicing TWAS. **e.** 3′aTWAS genes overlap in different cancer types. **f.** Heatmap showing the 3′aTWAS genes shared across different cancer types. The color represents the *P*-values for 3′aTWAS results. **g.** Effect of cancer-susceptibility-associated APA-linked genes on cell cancer proliferation based on experimental data from DepMap. Red dashes denote the median CERES cutoff value of < –0.5, which indicates an essential role in cell proliferation.

We then analyzed the identified genes across cancer types and found that breast and prostate cancers have the greatest number of significant APA-linked susceptibility genes (Fig. 4d). In addition, many of these genes are shared across multiple cancer types (Fig. 4e-f), such as the pan-cancer 3′aTWAS gene sorting nexin 17 (*SNX17)*, which plays a role in signaling-receptor and phosphatidylinositol binding, and is associated with breast and lung cancer^37^. Similarly, the 3′aTWAS gene spermatogenesis-associated 33 (*SPATA33),* which encodes a mammalian germline mitophagy receptor, is simultaneously related to the risk of melanoma, basal cell carcinoma, and skin cancer. Three APA genes common to breast and skin cancer were also identified, including the known tumor suppressor gene *CASP8*, a central mediator of the extrinsic apoptosis and necroptosis pathways^38^ (Fig. 4f).

To further assess the functional roles of putative APA-linked cancer susceptibility genes, we determined the effect of gene silencing on proliferation in cancer-relevant cell lines based on data from existing Clustered Regularly Interspaced Short Palindromic Repeat (CRISPR)–Cas9 screens. At a cutoff of median CERES score < –0.5^39^, our results indicate that numerous APA-linked genes play essential roles in cell proliferation (Fig. 4g). Notably, multiple known susceptibility genes with evidence for essential roles in breast cancer cell proliferation, such as *NUP107,* were found. In addition, we identified several novel putative susceptibility genes linked to prostate cancer cell proliferation, including *TUBA1B*, *EIF3E,* and *SUPT4H1*. These data indicate that APA might play a dominant role in regulating transcriptomic heterogeneity in multiple cancers.

### Identification of several novel cancer susceptibility APA genes significantly alters cell proliferation

Our colocalization and 3′aTWAS results identified multiple novel APA-associated cancer-susceptibility genes. To functionally assess their roles in tumor cellular phenotypes, we selected four putative breast cancer susceptibility genes—autocrine motility factor receptor (*AMFR*), cardiolipin synthase 1 (*CRLS1*), RPA-interacting protein (*RPAIN*), autophagy related 10 (*ATG10*)*—*that were identified by both colocalization and 3′aTWAS for further experimental validation. Importantly, besides *ATG10,* these candidates were not identified by expression or splicing TWAS. As examples, we show the LocusZoom and coverage plots for the 3′aTWAS genes *CRLS1* (Fig. 5a, c) and *RPAIN* (Fig. 5b, d). When conditioned on the 3′aTWAS association, the associated breast cancer signals substantially decrease (Fig. 5e, f), which indicates that our 3′aTWAS association can largely explain the GWAS signal in this region. Positive Z-scores of *CRLS1* and *RPAIN* in breast mammary tissue indicate that long 3′UTR of these genes increases the breast cancer risk. Although truncation of the 3′UTR is more commonly observed in cancers, genes with the elongated transcript can also enhance tumorigenesis^40^.

**Figure 5.**
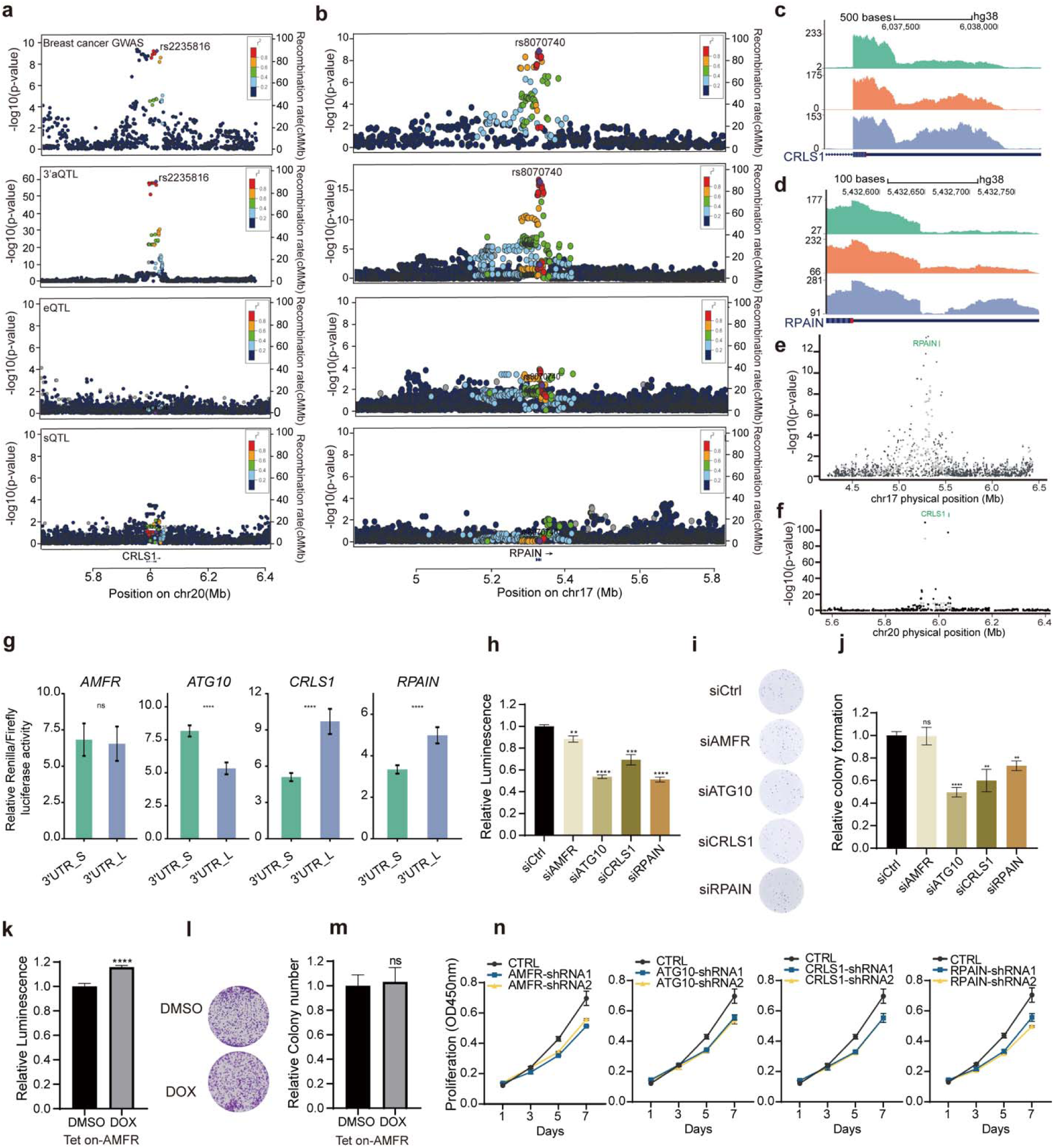
APA-linked susceptibility genes in breast cancer. **a,b.** LocusZoom plot of breast cancer GWAS SNPs, eQTLs, sQTLs, and 3′aQTLs at the (a) *CRLS1* and (b) *RPAIN* locus. SNPs are colored by LD (*r^2^*). **c,d.** Coverage plots illustrate that the respective breast-cancer-risk variants are strongly correlated with changes in the 3′UTR of (c) *CRLS1* and (d) *RPAIN* for each genotype. **e,f.** Region association plots for (e) *RPAIN* and (f) *CRLS1*. Breast cancer GWAS signal at the gene locus (gray) and the GWAS signal after removing the effects of target gene 3′UTR usage (black); results indicate that the association is largely explained by target gene 3′UTR usage. **g.** Luciferase activity from a reporter system containing the short and long 3′UTR of each breast cancer candidate gene in MCF-7. **h**. Proliferation of MCF-7 cells treated with the indicated small-interfering (si)RNAs. **i,** Colony formation assay for MCF-7 cells treated with the indicated siRNAs against breast cancer target genes. Colonies were formed in 12-well plates and imaged on day 8 after siRNA treatment. Images are representative results from three independent experiments. **j,** Quantification of colony numbers from panel (i). **k.** Proliferation of MCF-7 cells treated with DOX (1 μg/mL) or dimethyl sulfoxide (DMSO) control. **l,** Colony formation assay with MCF-7 cells treated with DOX (1 μg/mL) or DMSO control. Colonies were formed in 12-well plates and imaged on day 8 after treatment. Images are representative results from three independent experiments. **m,** Quantification of colony numbers in panel (l). **n**, Cell proliferation of shRNA mediated knockdown cells was analyzed at 1, 3, 5 and 7 days. For panels **h,j** and **k,m** the bar graphs show the mean ± standard deviation values from three independent experiments. Significance was determined by the Student′s *t*-test (two-tailed, unpaired), comparing the experimental group with the control. **, *P* < 0.01; \*\*\**P* < 0.001; ****, *P*<0.0001; ns, not significant.

Here, to further investigate the role of poly(A) site usage in regulating candidate gene expression, we inserted the shorter 3′UTR and the longer 3′UTR with a mutated proximal polyadenylation signal (PAS) into a dual-luciferase reporter system. For *RPAIN* and *CRLS1*, we found that the reporter containing the longer 3′UTR exhibits significantly higher luciferase activity than the construct containing the short 3′UTR in breast cancer cells MCF-7(Fig. 5g). These data imply that the longer 3′UTR promotes elevated protein levels, which increase breast cancer risk.

Based on the above observations, we hypothesized that these genes may possess oncogenic activities. To further test this possibility, we performed gene knockdown experiments with small-interfering (si)RNAs targeting each of the four candidate genes. All genes were expressed in human breast cancer cells MCF-7. Knockdown efficiency was calculated for each siRNA pair, and robust knockdown of the gene of interest was validated by quantitative reverse transcription (qRT)-PCR (Fig. S3). We then performed cell growth assays to evaluate cell survival and proliferative ability following gene knockdown (Fig. 5h). Results show that knockdown of *CRLS1, RPAIN,* and *ATG10* results in a significant reduction in cell proliferation in both cell lines. We also evaluated the effect of gene knockdown on colony-forming ability in MCF7 cells and found that knockdown of *CRLS1, RPAIN,* and *ATG10* significantly decreases colony formation compared to the non-targeting control (NTC) (Fig. 5i, j). Although we observed slight but insignificant changes in colony formation for cells with *AMFR* knockdown, both knockdown and overexpression of *AMFR* have significant effects on cell proliferation, indicating a possible role in cell activity and metabolism (Fig. 5l, m). Moreover, downregulation of *CRLS1, RPAIN, ATG10, and AMFR* via lentivirus-mediated short hairpin (shRNA) also significantly decreased breast cancer cell proliferation (Fig. 5n, Fig. S3). Thus, we have experimentally validated the roles of four APA-associated breast cancer susceptibility genes in tumor cell proliferation.

### Known and novel cancer susceptibility APA genes form coherent functional pathways

We hypothesized that the novel genes identified from colocalization, summary-data-based Mendelian randomization (SMR), and 3′TWAS may be connected to known cancer susceptibility genes via the same network or pathway. We then investigated whether our APA cancer-susceptibility genes are also enriched in public cancer-related susceptibility gene sets from the Molecular Signatures Database (MGB)^41^, DORGE^42^ and Cancer Gene Census (CGC)^43^. Notably, we identified 63 genes with well-established roles in cancer biology, such as predisposition genes for breast cancer (*ADNP, ALG12, CASP8, CDT1, COG7, CYP21A2, DHX37, EXOC2, FGFR2, HS2ST1, KANSL, NELFA, NF1, NUP107, PIGG, PIGN, RBM28, SMPD4*), basal cell carcinoma (*CASP10)*, cancer of female genital organs (*ATM, BPTF, HRAS, MSX1, STN1, WRAP53*), skin cancer (*ANKRD11, CASP10, CASP8, CDH3, KANSL1, MC1R, RFX5*), cancer of male genital organs (*KANSL1, RTEL1, WDPCP*), and lymphoma (*HLA-DQA1, HLA-DRB1*). Further, we used STRING^44^ to measure protein–protein interaction (PPI) network connectivity between the products of genes identified in this study and known cancer-susceptibility proteins. Results show that members of the 3′aTWAS-prioritized PPI network are highly enriched for key biological pathways (Fig. 6), including the superoxide metabolic process (*P* = 5.42 × 10^-3^), which play important regulatory functions in cancer cells′ reprogrammed metabolism, signaling and transcription^45^, and intracellular protein transport (*P* = 2.13 × 10^-2^) that is central to metabolic reprogramming during cancer progression and adaptation to new environment ^46^. More importantly, we observed that APA susceptibility genes strongly enriched in pathways related to apoptosis and necrosis, such as the execution phase of apoptosis with *P* = 5.88×10^-3^, TRAIL signaling with *P* = 7.47×10^-4^, which is essential to tumorigenesis ^47^. These results indicate that the novel candidate genes identified here function within more extensive gene networks having discrete biological functions. In particular, we show that our novel cancer-susceptibility genes form coherent functional pathways with known cancer genes, suggesting that many of these novel genes are likely causal.

**Figure 6.**
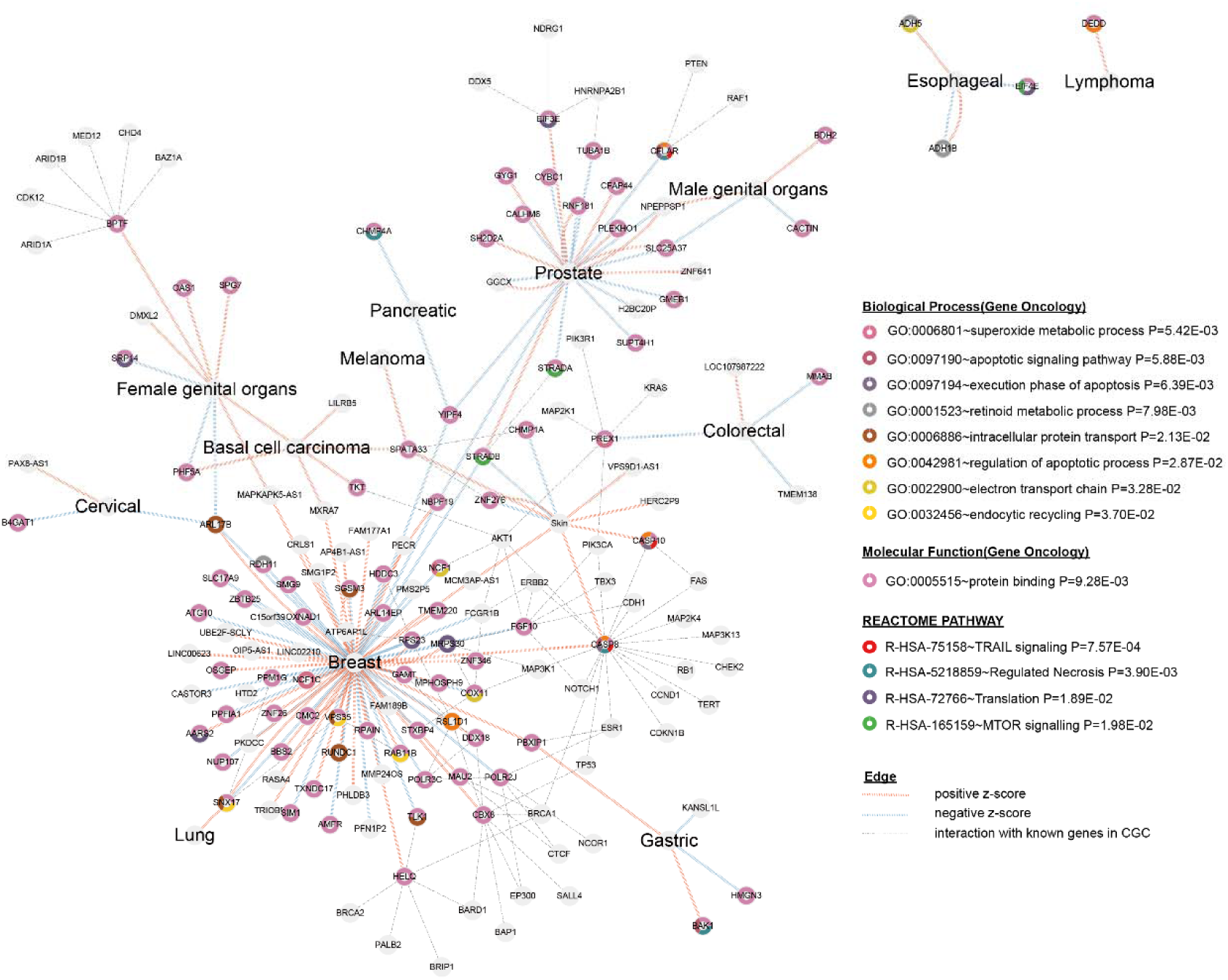
Newly identified APA-linked cancer susceptibility genes are enriched in metabolic reprogramming and apoptosis related pathways. APA-linked genes are connected in protein– protein interaction (PPI) networks with known cancer-risk genes, such as *CASP8* in breast and skin cancer. Pathway enrichment analysis shows that 3′aTWAS genes are enriched in cancer-related pathways, including the metabolic reprogramming pathway like superoxide metabolic process (*P* = 5.42 × 10^-3^) and intracellular protein transport (*P* = 0.021), the apoptosis related pathway like apoptosis signaling pathway (*P* =5.88 × 10^-3^) and TRAIL signaling pathway (*P* =7.57 × 10^-4^). Each node represents one APA-linked gene. Blue links between nodes represent a negative Z-score in 3′aTWAS, implying that usage of the short 3′UTR increases cancer risk, whereas red lines represent positive z-scores, indicating that usage of the longer 3′UTR leads to increased cancer risk. Gray dashed line indicated the PPI interactions between APA genes and the known cancer genes in CGC.

## Discussion

Here, we performed the first comprehensive study evaluating the genetic role of APA across human cancer types. We demonstrate that 96.25% of leading cancer-associated SNPs are noncoding and 14.1% were located in 3′UTR and downstream 20kb regions. We also found that fine-mapped SNPs showed strong enrichment in noncoding regions, whereas they are depleted in the intronic region. By leveraging the largest compilation of cancer summary statistics and large-scale transcriptomic data from 49 tissues, we further identified 413 cancer susceptibility genes linked to APA. Our findings identify APA as an important mechanism to explain the function of cancer-associated noncoding variants, and therefore imply that future studies should consider APA as an intermediate molecular phenotype, which may contribute to disease risk.

This work advances our understanding of cancer-risk SNPs in a genomic post-transcriptional context. Moreover, these findings add to a growing body of evidence demonstrating that APA is a general mechanism for post-transcriptional gene regulation during various physiological states and in complex diseases. Critically, genes may be associated with cancer through their molecular function or via interaction with causal genes. Here, to determine the potential functional significance of our APA-linked genes and provide evidence for causal inference, we performed knockdown studies with four genes for which increased expression is associated with poor cancer prognosis. We then measured the effect of knockdown on proliferation and colony-forming efficiency in breast cancer cell line, observing the strongest and most consistent effects in cells treated with siRNA targeting *CRLS1* and *RPAIN*. Thus, further experimental study is necessary to evaluate the functions of these and other APA-linked cancer-risk genes and elucidate the mechanisms by which they may modulate downstream protein expression and risk of disease development in cancer.

Our study prioritizes multiple gene candidates for subsequent experimental follow-up, focused on interrogating the molecular mechanisms underlying cancer progression and development. Critically, further investigation of these genes identified in our study has the potential to provide new mechanistic insights into the pathology and genetics of various cancers. We also provide a valuable resource (http://bioinfo.szbl.ac.cn/TCGD/index.php), which provides an important resource for the community to interpret the function of cancer risk variants and offers beneficial resources for exploring the genetic basis in a wide variety of human cancers.

## Materials and Methods

### Sources of GWAS summary statistics

We collected and integrated GWAS summary statistics for a total of 33 tumor types from three different sources: (1) summary statistics reported in the literature, including the NHGRI–EBI GWAS Catalog^23^, GWAS ATLAS, JENGER, and published studies; (2) publicly available summary statistics from the UK Biobank Imputed Dataset v.3^24^; and (3) combined genome information and digital health care data from national health registries of the FinnGen Biobank. Finally, CrossMap^48^ was used to convert the coordinates of the GWAS summary statistics to the human genome assembly hg19/GRCh37.

#### Reported GWAS

We downloaded previously reported GWAS variants from various public repositories. The first set of reported GWAS summary statistics was based on 4,795 publications and 222,481 associations (the NHGRI–EBI GWAS Catalog, file version: 2020-12-02). The data were mapped to Genome Assembly GRCh38.p13 and dbSNP Build 153. The second set of reported GWAS summary statistics contained 4,756 statistics from 473 unique studies across 3,302 unique traits and 28 domains. Among them, 600 GWASs in this project were performed based on UK Biobank Imputed Dataset (v3). Therefore, this part of the data is partially duplicated with the “round 2” version of the UK Biobank-based GWAS. The third set of reported GWAS summary statistics was based on the JENGER from BioBank Japan (BBJ). In addition, we also extracted partial GWAS summary statistics from recent publications.

#### UK Biobank-based GWAS

We downloaded UK Biobank GWAS summary statistics from a public repository (UKB GWAS Neale lab). The updated “round 2” version of UK Biobank GWAS data expands the number of phenotypes to 4,203 (up from 2,419 in round 1), included more individuals (*i.e*., 361,194), and by popular demand, generated sex-specific results to accompany the main GWAS results.

#### FinnGen GWAS

We downloaded Finnish genome information and digital health care data from the FinnGen (FinnGen FREEZE 4, update on Nov 30, 2020). This newest update contains data from 176,899 individuals, almost 17 million variants, and 2,444 disease endpoints. As a result, we downloaded 173 groups of cancer-associated GWAS summary statistics.

### Processing of GWAS summary statistic data

Publicly available GWAS summary statistics were curated from multiple resources and included only when the full set of SNPs was available (last update January 20, 2021). Curated summary statistics were pre-processed to standardize the format. SNPs with *P* ≤ 0 or > 1 or non-numeric values such as “NA” were excluded. For summary statistics with non-hg19 genome coordinates, CrossMap^48^ software was used to align to hg19. Genome coordinates were extracted from dbSNP 148 only when an rsID was available in the summary statistics file without chromosome and position. When the rsID was missing, it was assigned based on dbSNP 148. When only the effect allele was reported, the other allele was extracted from dbSNP 148.

### Definition of leading SNPs and cancer-associated loci

For each GWAS, leading SNPs and genomic trait-associated loci were identified, as described previously. First, we defined significant independent SNPs with *P* < 5 × 10^−8^ and independence at *r^2^* < 0.6 and identified linkage disequilibrium (LD) blocks based on SNPs with *P* < 0.05. Of these SNPs, we further defined leading SNPs that are independent at *r^2^* < 0.1. We finally determined cancer-associated loci by merging LD blocks closer than 250 kb. Each trait-associated locus was then represented by the top SNP (with the minimum *P*-value), and its genomic region was defined by the minimum and maximum position of the significant independent SNPs within the merged locus with an LD (*r^2^* ≥ 0.6). SNPs with minor allele frequency (MAF) < 0.01 based on the summary data were excluded from all analyses due to lower statistical power and a high false-positive rate among SNPs with extremely small MAFs.

We used the 1000 Genome Project phase 3 (1000G3) as a reference panel to compute LD for most GWASs in the database. For each GWAS, a matched population—comprising African (AFR), American (AMR), East Asian (EAS), European (EUR), or South Asian (SAS) individuals—was used as the reference based on information For trans-ethnic GWASs, the population with the largest total sample size was used. For GWAS based on the UK Biobank release 1 cohort (UKB1), we used 10,000 randomly sampled unrelated white British subjects from UKB1 as a reference. For GWASs based on the UKB2, 10,000 randomly selected unrelated EUR subjects were used as a reference. The reference panel for each GWAS is specified in Supplementary Table 3.

### Estimated standardized effect size of leading SNPs

Leading SNPs were defined using a genome-wide significance cutoff of *P* < 1×10^-8^ and an *r^2^* of 0.1, based on the population-relevant reference panel. To enable comparison of effect size across different studies, regardless of the direction of effect, we converted *P*-values into Z-statistics (two-sided) and expressed the standardized effect size (β) as a function of MAF and sample size, as described previously, using the following equation:

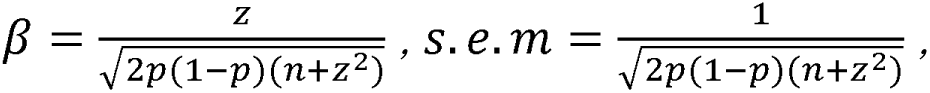

where *p* is MAF, and *n* is the total sample size. We used the MAF of a corresponding European reference panel (either 1000 genome phase 3, UKB1, or UKB2), as described above in ‘Definition of leading SNPs and trait-associated loci’.

### Fine mapping of GWAS

CAUSALdb^25^, which includes three fine-mapping methods (PAINTOR, CAVIARBF, and FINEMAP), was applied to the GWAS summary statistics data to identify causal SNPs. LD information of GWAS variants was estimated in each LD block using five super-populations (AFR, AMR, EAS, EUR, and SAS) from the 1000 Genome phase 1 (1000G1) reference panel. Because Ldetect^49^ only contains LD block information for three continental populations, the four 1000G1 super-populations were assigned to these three populations by mapping EUR and AMR to the European population, EAS to the Asian population, and AFR to the African population. We then defined SNPs belonging to 95% credible sets as causal by fine mapping, allowing only one causal SNP for each LD block, using the recommended tool parameters. We further note that FINEMAP requires variants MAFs, and when this information was unavailable in the original file, we used data in the reference panel.

### SNP genome-wide heritability and genetic correlations

LDSC^26^ on summary statistics from the union set of all SNPs was used to estimate the genome-wide liability-scale heritability of each cancer type and the genetic correlation between each pair of cancer types. Prepared LD scores for 1000 genome phase 3 EUR and EAS populations were obtained from https://data.broadinstitute.org/alkesgroup/LDSCORE/, and LDSC was performed for GWASs based on either a EUR or EAS population, respectively; the number of SNPs in all summary statistics files was required to be >450,000. SNPs were limited to those in HapMap3, with the major histocompatibility (MHC) region (25–34 Mb) excluded. When the signed effect size or odds ratio was not available in the summary statistics, an ‘—a1 -inc’ flag was used. As recommended in the literature, we removed SNPs with χ^2^ > 80^50^. Genetic correlations were computed for pairwise GWASs with the following criteria, as described previously^50^: (1) GWASs of either EUR or EAS populations with > 80% of samples having corresponding ethnicities; (2) number of SNPs totaling >450,000; (3) signed statistics available. The *r_g_*estimated by LDSC is an unbiased estimate and may exceed [-1,1] when standard errors are large, and the genetic correlation between studies is high. Genetic correlations for which the *P*-value survived the correction for multiple testing, with Bonferroni-corrected *P* < 0.05, were considered significant.

### Test enrichment of GWAS signals in xQTLs

To assess the GWAS–xQTL enrichment and test whether a set of xQTLs in a given tissue is enriched for subthreshold-(for example, 5 × 10^-8^ < *P* < 0.05) to genome-wide-significant (*P* ≤ 5 × 10^-8^) common variant associations with a given complex disease or trait (*i.e*., more than would be expected by chance) we used *fgwas*. To this end, we extracted GWAS SNPs also belonging to xQTLs (association *P* < 10^-4^) and plotted the QQ-plot of the GWAS *P*-values for those SNPs. Genome-wide GWAS *P*-values were also plotted as a control.

## Quantification of GTEx v.8 APA levels using DaPars2

Our previously developed DaPars2 software^51^, which allows the joint analyses of multiple samples based on a two–normal mixture model, was used to calculate the poly(A) site-usage index (PDUI) value. Briefly, we extracted a 3′UTR annotation for each gene using the “DaPars_Extract_Anno.py” script within DaPars2. We then used the “samtools flagstat” command to calculate the sequencing depth for each sample. Finally, we used DaPars2 to calculate the PDUI value of each transcript across samples.

### 3′aQTL mapping

As previously described^21^, we applied a linear regression framework in Matrix eQTL^52^ to test the association between normalized PDUI values and SNPs within an interval of 1 Mb from the 3′UTR region, adjusting for known covariates (including sex, RIN, platform, and top-five genotype principle components) and unobserved covariates calculated by PEER^53^. The number of PEER covariates for each tissue was as suggested by the GTEx Consortium. We performed 1,000 rounds of permutation to obtain empirical *P*-values for each gene, which were then adjusted using the R package qvalue.

### Colocalization tests

We first downloaded all variant–gene cis-eQTL and cis-sQTL associations tested in each tissue data from GTEx v8. We then constructed all variant–transcript cis-3′aQTL associations tested in each tissue data from GTEx v8. To restrict the total number of intended colocalization tests to a computationally tractable number, we first performed an I overlap test of the GWAS summary statistics and eQTL, sQTL, and 3′aQTL association summary statistics from GTEx v.8. For each GWAS, we selected all SNPs with a nominal association *P* < 5 × 10^-8^, chosen as the least stringent threshold that was computationally tractable. We also required that selected SNPs be at least 1-Mb apart from all SNPs already selected from the same GWAS, to ensure independence of effects and distinct loci. For every selected GWAS SNP, we identified all eQTL, sQTL, or 3′aQTL features (*i.e*., gene expression, splice junction usage, and 3′UTR usage variant, respectively) with a QTL association *P* < 1 × 10^-5^ at any SNP positioned within 100 kb of the most significant GWAS SNP at the locus. We made no quantitative claims about the significance of these naïve overlaps, given that many such overlaps could be expected by chance; these merely formed the set of loci to test for colocalization in subsequent steps.

To identify the typical genetic effects between a given GWAS trait and a given QTL, we used a Bayesian colocalization method within the *coloc* v.4.0.4 R package^54^. For each cancer GWAS trait, we extracted the sentinel SNP, defined as a GWAS SNP with *P* < 5 × 10^−8^, having the greatest significance within at least 1 Mb. We then searched for colocalized QTL signals within the 100-kb region surrounding each sentinel SNP. As defined by the *coloc* method, five posterior probabilities (PPs) were calculated: PP0, null model of no association; PP_1_, only the GWAS SNP has a genetic association in the region; PP_2_, only the QTL has a genetic association in the region; PP_3_, both the GWAS SNP and QTL are associated, but with different causal variants; PP_4_, both the GWAS SNP and QTL are associated and share a single causal variant. The genes were defined as colocalization events if PP_4_L≥L0.75 and PP4/(PP4L+LPP3)L≥L0.9. Region visualization plots were constructed using LocusZoom v.1.4^55^. LDs between reference SNPs and 3′aQTLs were calculated using PLINK^56^.

For comparison, we also performed SMR^11^ analysis. In brief, SMR applies the principles of Mendelian randomization (MR) to integrate summary statistics from GWAS and xQTL data, in order to test for associations between features (*e.g*., gene expression, splicing, and APA) and traits and prioritize genes underlying GWAS associations. Here, variant–feature and variant–trait association statistics in a two-sample MR framework were used in SMR tests to identify associations between features and traits. The heterogeneity in dependent instruments (HEIDI) test, typically applied to significant SMR results, eliminates cases where the association is driven by linkage or proximity of independent causal variants rather than a shared causal variant. We ran SMR using the default parameter settings to obtain an SMR *P*-value for each locus and a HEIDI *P*-value for each locus.

### APA TWAS for cancer GWAS

We used FUSION^35^ to perform TWAS for APA. Briefly, for each reference panel, FUSION estimates the heritability of 3′UTR usages explained by local SNPs within a 1-Mb region proximal to the 3′UTR of each transcript using linear-mixed models. Known covariates, such as sex and genotyping platform, and hidden batch effects or other unobserved covariates from PEER were included to determine residualized PDUI values calculated by DaPars2. Residualized PDUI values were utilized to train cross-tissue 3′aTWAS models with genotype data, and prediction models with a heritability of Bonferroni-corrected *P* < 0.05 were used for further analysis (Fig. S1). The 3′UTR predictive weights were computed by five different models implemented in the FUSION framework: BLUP, LASSO, Elastic Net, and top SNPs. Cross-validation for each of the desired models was then performed.

For comparison, S-PrediXcan^57^ was employed to incorporate different cancer GWAS data and expression and splicing QTL information from each of the 49 GTEx tissues (v.8) and identify potential risk genes associated with different cancer types. We obtained gene expression and splicing pre-specified mashr weights based on covariance of genetic variants from the PredictDB data repository (http://predictdb.org), and those weights were used in S-PrediXcan analysis.

Lastly, the imputed feature usages (*i.e*., expression, splicing, and APA) are correlated with cancer GWAS summary statistics to perform TWAS and identify significant associations. To account for multiple hypotheses, we applied an FDR of 0.05 within each feature (*i.e*., expression, splicing, and APA) reference panel.

### Joint conditional probability (JCP) analysis

We often identify multiple associated features in a locus (or the same feature from multiple tissues). Hence, it is necessary to identify which loci are conditionally independent. We therefore performed JCP testing for 3′aTWAS significant associations (FDR < 0.05) to assess the independence of associations within their respective 1-Mb windows. JCP analysis was run on all candidate hit regions with FUSION.post_process.R to disentangle signals within regions containing multiple significant genes by examining the probability that multiple associations occur simultaneously (jointly). This analysis module helps to identify genes that are associated, irrespective of surrounding genes (Marginal), from those that rely on surrounding loci (Conditional).

### Annotation of 3′TWAS-identified genes in cancer-relevant gene databases

We collected cancer-related gene sets from the Molecular Signatures Database (MSigDB to identify overlap between our 3′TWAS-identified genes and known cancer susceptibility genes. Putative cancer-related genes were identified by annotating with specific key phrases, such as “breast cancer” and “prostate cancer”. Among the APA-associated identified genes in our investigation, we determined the number of probable cancer-related genes that overlap with those obtained from the MSigDB^41^. We also determined whether the genes identified in this study were overrepresented in the set of DORGE^42^, and CGC genes^43^ from the COSMIC website (https://cancer.sanger.ac.uk/census).

### Effects of CRISPR–Cas9 gene silencing on proliferation in cancer-relevant cells

Gene-dependency levels for 17,386 genes based on CRISPR-Cas9 essentiality screen data, as determined by the CERES computational method, were downloaded from the DepMap portal^58^. For each gene, we calculated the total count and median of negative CERES values (for cell proliferation) from relevant cancer cells. A cutoff CERES value < −0.5 was used to determine essentiality.

### Dual-luciferase reporter assay

Renilla luciferase in psiCHECK-2 Vector (Promega, cat. no. C8021) was used as a primary reporter gene. The short 3′UTR and long 3′UTR with mutated proximal PAS sequence of *AMFR*, *CRLS1*, *RPAIN*, *ATG10* was cloned downstream of the Renilla luciferase translational stop codon of psiCHECK-2 Vector, respectively. The firefly reporter cassette served as an intra-plasmid transfection normalization reporter^59^.

For dual-luciferase reporter assay, MCF-7 cells were seeded in 1 day prior to transfection. The previously described luciferase psiCHECK-2 vector (constructed by Tsingke biotech and confirmed by sequencing) were transfected into cells using Lipofectamine 3000 Transfection Reagent (Invitrogen, cat# : L3000015) according to the manufacturer′s instructions. Forty-eight hours post-transfection, firefly and renilla luciferase activities were measured by Dual-Luciferase Assay System (Promega, #E1980) on a BioTek Synergy H1 plate reader with full waveband. Each assay was measured in three independent replicates.

### Gene knockdown with siRNA and quantitative RT-PCR

MCF-7 cell lines obtained from the Cell Resource Center of Shanghai Institutes for Biological Sciences (Chinese Academy Science, Shanghai, China)^11^ were cultured in Dulbecco’s Modified Eagle Medium (DMEM; cat #: C11995500BT; Gibco, Thermo Fisher Scientific, Waltham, MA, USA), containing 10% fetal bovine serum (FBS; Gibco, cat #: C10010500BT), 100-IU/mL penicillin, and 100-μg/mL streptomycin (Gibco, cat #: 15140-122). Cells were cultured at 37°C in a 5% CO_2_ atmosphere with 100% humidity. When cells reached 60–80% confluency, they were transfected with NTC siRNA or a pool of three different gene-specific siRNAs (**Table S7**; RiboBio, Guangzhou, Guangdong, China^43^), using Lipofectamine RNAiMAX Reagent (cat #: 13778150; Thermo Fisher Scientific), at a final concentration of 50 nM, according to the manufacturer′s protocol. The medium was replaced after 12 h, and cells were harvested 48 h after transfection.

Total RNA was extracted using the Quick-RNA™ Miniprep Kit (cat #: R1055; Zymo Research, Irvine, CA, USA), and cDNA was generated using the Hifair® L 1st Strand cDNA Synthesis Super Mix for qPCR (gDNA digester plus) kit (cat #: 11141ES60; YEASEN, Shanghai, Shanghai, China). Quantitative PCR was performed using the Hieff® qPCR SYBR Green Master Mix (cat #: 11203ES08; YEASEN) on a CFX96 machine (BIO-RAD, Hercules, CA, USA). Primers used for qPCR are listed in **Table S8**. Experiments measuring expression of each gene were repeated at least three times, with *GAPDH* used as the internal reference for expression.

### Cell proliferation assay

Cell proliferation assays were performed using the CellTiter-Glo 2.0 Kit (cat #: 92243; Promega, Madison, WI, USA). Briefly, MCF-7 cells were transfected with siRNA for 16–20 h. A total of 5,000 cells from each treatment were then re-plated into 96-well plates in quadruplicates. After 72 h, the medium was replaced with 200 μL of a 1:1 mixture of DMEM and CellTiter-Glo 2.0 reagent. Doxycycline-inducible AMFR MCF7 cells were used for AMFR overexpression, 1000 cells were seeded into 96-well plates in quadruplicates. After 24 hours, cells were treated with doxycycline (Sangon Biotech, cat#: A600504-0025) at a final concentration of 1ug/mL. After 6 days, the medium was replaced with 200 μL of a 1:1 mixture of DMEM and CellTiter-Glo 2.0 reagent. The cells were lysed on an orbital shaker at 300 rpm for 2 min at room temperature. The plates were equilibrated for 10 min, and the luminescent signal, reported as relative light units (RLU), proportional to the amount of ATP, was measured at the full waveband with a BioTek Synergy H1 plate reader. Results were calculated by GraphPad Prism v.8.0.1 (GraphPad, San Diego, CA, USA).

### Cell colony formation assay

MCF-7 cells were transfected with siRNA for 16–20 h, and 2,000 cells were then re-plated into 12-well plates in triplicates for each gene. A total of 1000 cells from doxycycline-inducible AMFR MCF-7 cell line were seeded into 12-well plate in triplicates. After 24 hours, cells were treated with doxycycline (1 μg/mL). When single clones contained more than 50 cells, the colonies were fixed with 4% paraformaldehyde for 20 min and stained with 0.1% crystal violet (cat #: A600331-0025; Sangon Biotech, Shanghai, Shanghai, China) for 20 min at room temperature. After washing with water, the plate was air dried before imaging. Colony counting was performed using ImageJ 1.53r (National Institutes of Health, Bethesda, MD, USA).

### Generation of stable knockdown cell line using lentivirus delivered shRNA

Two set of shRNAs against each gene was used to cloned into pLKO.1-puro vector. shRNA-expressing lentivirus was produced with the third-generation packaging system in human embryonic kidney (HEK) 293T cells (ATCC, CRL-11268). Briefly, 70–80% confluent 293T cells in 6-well plate were transiently cotransfected with 5 μg of lentiviral transfer vector, 1.67 μg of pVSVG (envelope plasmid), 1.67 μg of pRSV-Rev (packaging plasmid) and 1.67 μg of pMDLg/pRRE (packaging plasmid) with PEI according to the manufacturer′s instructions. Medium was replaced 24 h after transfection with low-glucose DMEM GlutaMAX (Gibco) containing 10% FBS and 0.1% penicillin and streptomycin (Sigma), and virus supernatant was collected every 24 h for up to 2 d. Supernatent containing viral particles was filtered through a 0.45-μm filter unit (Millipore) and stored at −70 °C in aliquots or used directly for cell infection. For lentivirus infection, target cells were seeded in a 6-well plate or a 10-cm dish 16–18 h before infection and were grown to 70–80% confluency upon transduction. Culture medium was removed, and cells were incubated with virus supernatant along with 8 μg/ml polybrene (Sigma). After overnight incubation, virus-containing medium was replaced with fresh medium. Puromycin (Sigma) was applied to kill non-infected cells 36 to 48 h after infection. After 2 days of selection when non-infected control cells were all dead, surviving cells were split and maintained with the same concentration of puromycin. After 3 d, cells were collected for RNA and tested by RT-qPCR to confirm successful shRNA knockdown efficiency of target genes.

### Cell viability and proliferation assays for shRNA mediated knockdown

Cells were trypsinized, resuspended at 1 × 10^4^ cells/ml and seeded in 96-well plates with each well containing 100ul medium of 1 × 10^3^ cells. Cell viability and proliferation were determined with CCK8 assays (Yeasen, cat#: 40203ES76) at designed time points by reading the absorbance at 450 nm, following the manufacture′s instructions. Values were obtained from four replicate wells for each treatment and time point. Results are representative of three independent experiments.

### A comprehensive data portal to host cancer susceptibility genes

We constructed the Cancer GWAS Database (TCGD) (http://bioinfo.szbl.ac.cn/TCGD/index.php) as a comprehensive data portal for browsing, searching, and visualizing cancer susceptibility genes. TCGD has included 438 cancer GWAS summary statistics from 59 unique studies across 59 cancer types, which are provided for researchers and users to investigate the role of APA in human cancer. The users can explore cancer GWAS summary statistics in the “Browse GWAS” module by selecting cancer/neoplasm type, population type, published year, and total sample size. Meanwhile, all the columns in this summary table are allowed to search and sort in the demand of customers. In addition, they can list each GWAS with the Q-Q plot, Manhattan plot, risk loci and SNP heritability, and other detailed information in the link of the CGID. The multi-tissue TWAS data are organized in a tabular table as well. The users can browse and search their genes (e.g., *IRF5*) or information (e.g., 3′aQTL) for the multi-tissue TWAS. Then, the gene name can be clicked for browsing and searching the single-tissue TWAS.

## Data availability

Raw whole transcriptome and genome sequencing data from the Genotype-Tissue Expression (GTEx) project are available via the database of Genotypes and Phenotypes (dbGaP), under the accession number: phs000424.v8.p2^60^. All processed GTEx data are available via the GTEx portal((http://gtexportal.org/). GWAS summary statistics are from NHGRI-EBI GWAS catalog (https://www.ebi.ac.uk/gwas/), UK Biobank GWAS, http://www.nealelab.is/uk-biobank/), Finn Gen(https://www.finngen.fi/en) and JENGER(http://jenger.riken.jp). The details, including accession numbers, of GWAS summary statistics used in this study are listed in Supplementary Table 1. 1000 Genomes Project Reference for LDSC, https://data.broadinstitute.org/alkesgroup/LDSCORE/1000G_Phase3_plinkfiles.tgz; 1000 Genomes Project Reference with regression weights for LDSC, https://data.broadinstitute.org/alkesgroup/LDSCORE/1000G_Phase3_weights_hm3_no_MHC.tg z). All significant 3′aTWAS genes in cancer are available at Supplementary Table 6. The expression and splicing TWAS models for GTEx v8 are publicly available at PredictDB (https://predictdb.org/).

## Author Contributions

L.L., D. L., W.L. conceived and supervised the project. H.C., W.C., X.Z., performed the bioinformatics analysis. Z.W., J. W, C.L., performed the experiments. X.M., constructed the website. M. P., T.N., G-H, W contributed expertise in cancer and genetic analyses. H.C., Z.W., L.L. wrote the manuscript with the assistance from other authors.

## Data Availability

Raw whole transcriptome and genome sequencing data from the Genotype-Tissue Expression (GTEx) project are available via the database of Genotypes and Phenotypes (dbGaP), under the accession number: phs000424.v8.p260. All processed GTEx data are available via the GTEx portal((http://gtexportal.org/). GWAS summary statistics are from NHGRI-EBI GWAS catalog (https://www.ebi.ac.uk/gwas/), UK Biobank GWAS, http://www.nealelab.is/uk-biobank/), Finn Gen(https://www.finngen.fi/en) and JENGER(http://jenger.riken.jp). The details, including accession numbers, of GWAS summary statistics used in this study are listed in Supplementary Table 1. 1000 Genomes Project Reference for LDSC, https://data.broadinstitute.org/alkesgroup/LDSCORE/1000G_Phase3_plinkfiles.tgz; 1000 Genomes Project Reference with regression weights for LDSC, https://data.broadinstitute.org/alkesgroup/LDSCORE/1000G_Phase3_weights_hm3_no_MHC.tgz). All significant 3′aTWAS genes in cancer are available at Supplementary Table 6. The expression and splicing TWAS models for GTEx v8 are publicly available at PredictDB (https://predictdb.org/).

## Acknowledgments

We thank members of the Li laboratory for helpful discussions. This work was supported by the National Key Research and Development Program of China (no. 2022YFA1302800) to L.D., National Natural Science Foundation of China (no. 32100533) and Open grant funds from Shenzhen Bay Laboratory (no. SZBL2021080601001) to L.L. National Natural Science Foundation of China (no. 32270779) to L.D.. We also thank Qin Wang at Shenzhen Bay Laboratory supercomputing center for high-computing support.

## Supplementary Figures

**Figure S1.**
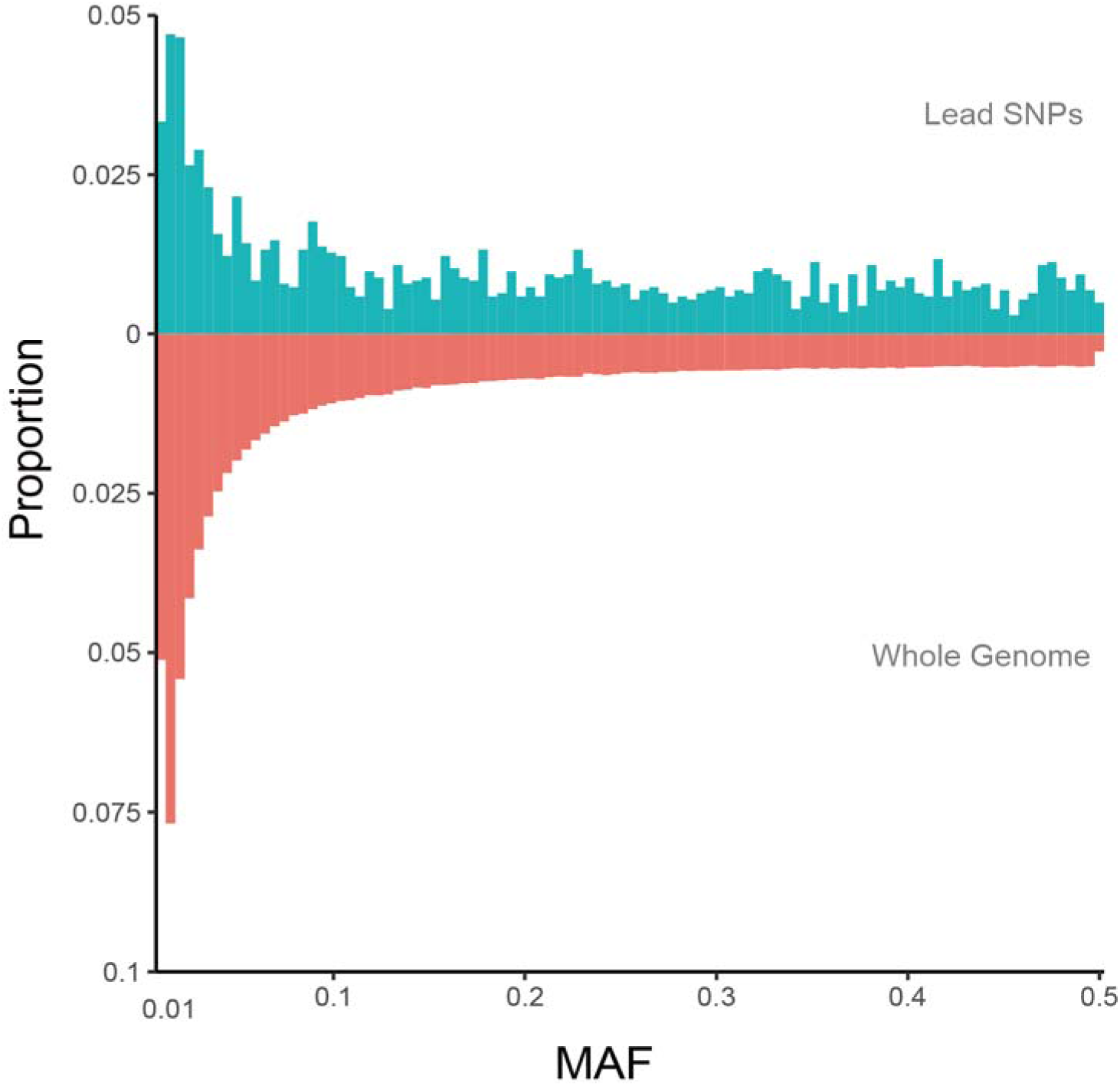
Histogram of minor allele frequencies (MAFs) for unique leading SNPs compared with the whole genome.

**Figure. S2.**
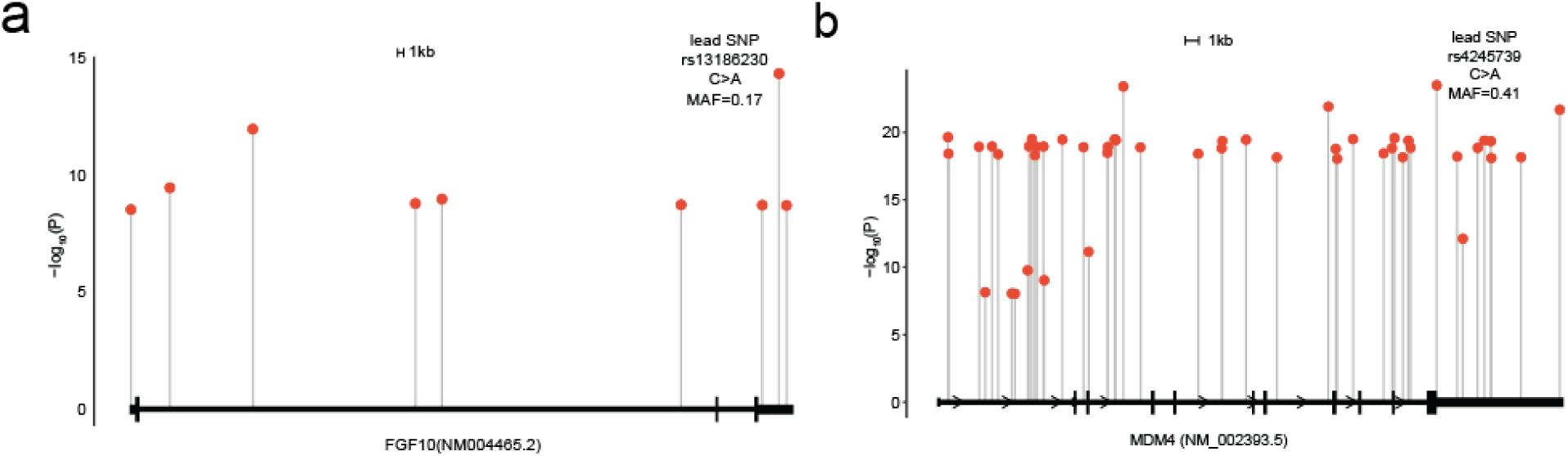
Examples of genes with lead cancer risk variants were located in 3′UTR regions.

**Figure S3.**
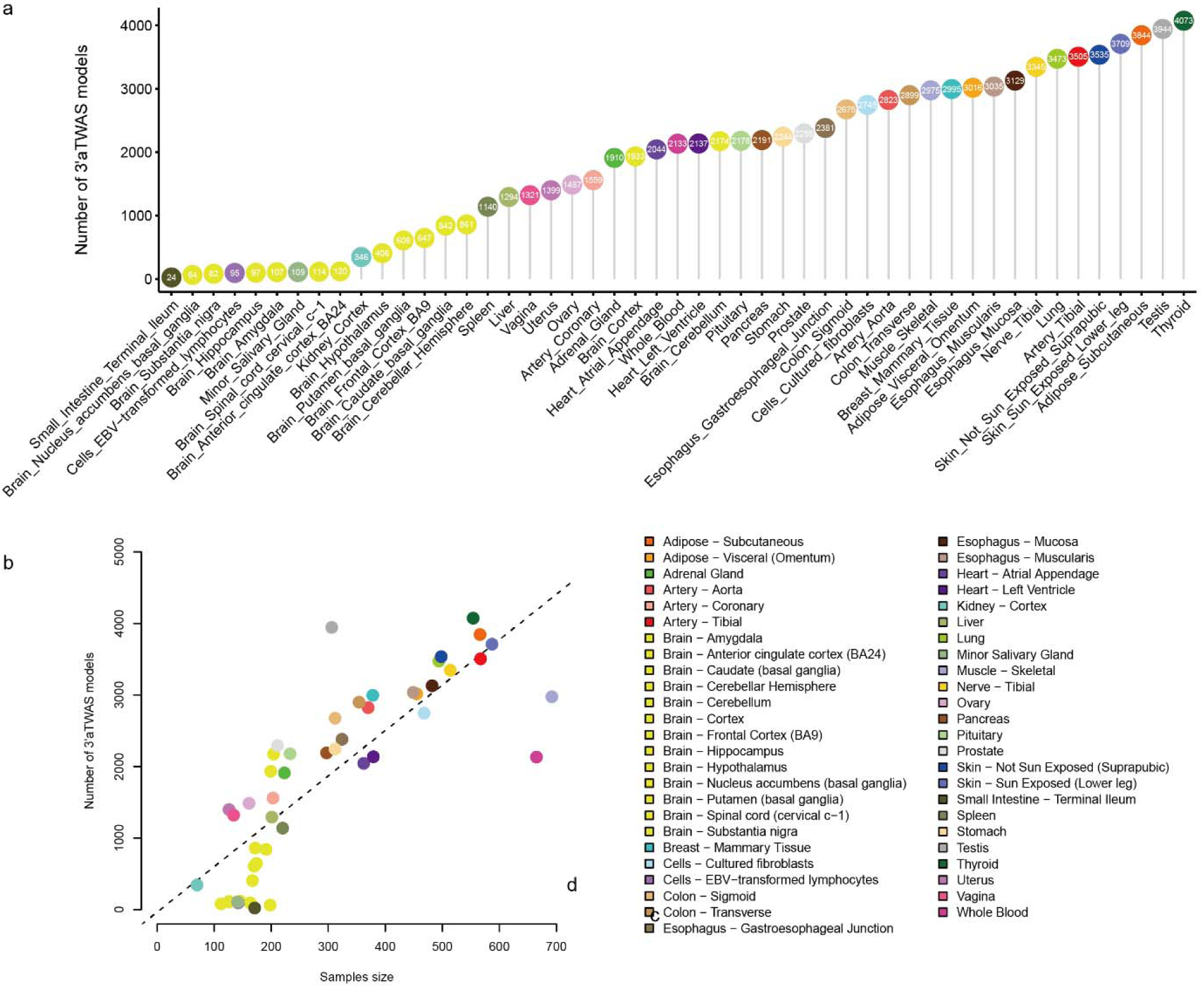
3′aTWAS models across 49 tissues from the Genotype–Tissue Expression (GTEx) cohort. **a.** Number of 3′aTWAS models across each tissue. **b.** The number of 3′aTWAS models is highly correlated with the sample size in each tissue. Each dot indicates a certain tissue type.

**Figure S4.**
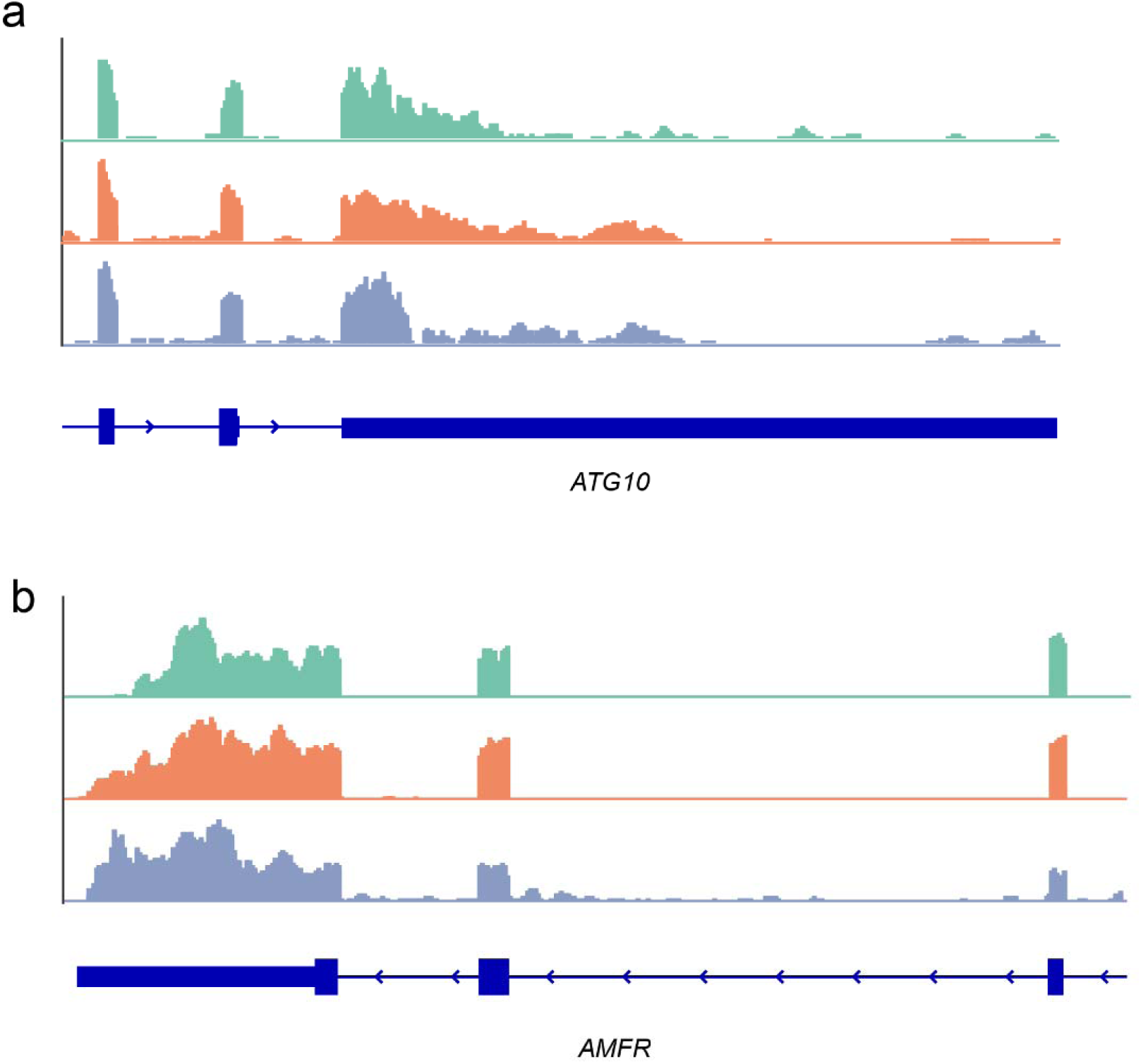
Coverage plot of *ATG10* and *AMFR* indicating the breast cancer risk variant correlated with 3′UTR changes.

**Figure S5.**
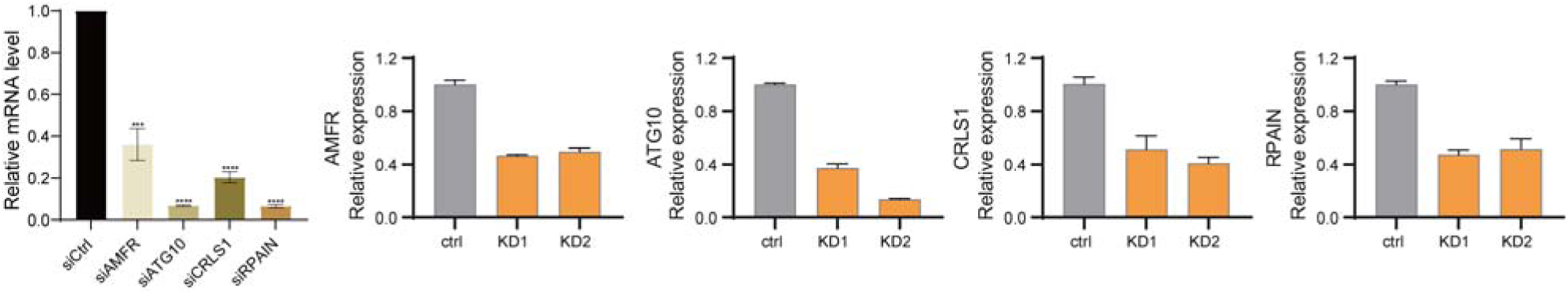
Validation of knockdown efficiency for the indicated siRNAs and shRNAs. Quantitative reverse transcription (qRT)-PCR measuring expression of the indicated genes in MCF-7. The bar graph represented mean ± standard deviation values from three independent experiments. Significance was determined by the Student′s *t*-test (two-tailed, unpaired), comparing siRNA-knockdown and siRNA control (siCtrl) cells; ***, *P* < 0.001; ****, *P* <0.0001.

